# Contextualizing the Utility of Polygenic Risk Scores using Absolute Risk Models in Diverse Ancestry Populations

**DOI:** 10.64898/2026.06.03.26354842

**Authors:** Martina Fu, Linda Kachuri, Dezheng Huo, John S. Witte, Pradeep Natarajan, Nilanjan Chatterjee

**Affiliations:** Department of Epidemiology and Population Health, Stanford University, USA; Department of Public Health Sciences, University of Chicago, USA; Department of Biomedical Data Sciences, Stanford University, USA; Department of Genetics, Stanford University, USA; Broad Institute of MIT and Harvard, Cambridge, MA, USA; Heart and Vascular Institute and Center for Genomic Medicine, Mass General Brigham, Boston, MA, USA; Department of Medicine, Harvard Medical School, Boston, MA, USA; Department of Biostatistics, The Johns Hopkins University, Baltimore, MD, USA; Department of Oncology, The Johns Hopkins University, Baltimore, MD, USA

## Abstract

Polygenic risk scores (PRSs) are emerging as powerful tools for quantifying inherited risk for common diseases and, in some cases, are approaching clinical implementation. A major concern for PRS implementation is their limited accuracy in non-European populations, particularly in those of African ancestry. However, past evaluations have focused on metrics such as relative risk or AUC, which do not capture background risk arising from contextual factors. We introduce a novel measure of variable importance, the conditional average derivative estimator (CADE), to evaluate PRS utility across diverse contexts and populations within absolute risk models that integrate PRSs with other relevant risk factors. We illustrate this framework by integrating PRSs for breast and prostate cancer within age-specific absolute risk models for incidence and mortality fit using individual-level data from the All of Us Research Program with inputs from the National Cancer Institute SEER cancer registry. Our projections show that although the PRSs are known to have the lowest discriminatory accuracy in African Americans (AA), there are contexts in which they provide greater utility, such as for the stratification of prostate cancer risk and mortality, where the CADE values for AA were 2- and 7-fold higher than for European Americans. These findings suggest that conclusions about the limited clinical utility of PRS in non-European populations may be premature and underscore the need to quantify PRS risk-stratification utility at the absolute-risk level, while accounting for disease onset, survival, and broader health and economic factors.

## Introduction

GWASs of increasingly large sample sizes and diverse populations have led to polygenic scores with substantial utility for risk stratification in many common diseases, such as breast^1^ and prostate cancers^2^, and coronary artery disease^3^. Various clinical studies are now investigating the utility of these scores in specific intervention contexts, such as for cancer screening^4,5^ and lifestyle modification^6^. Popular tools for risk prediction, such as the BOADICEA model for breast cancer, have been expanded to include polygenic risk scores^7^. Medical societies such as the American College of Cardiology and the American Heart Association have updated their guidelines to acknowledge PRSs as risk-enhancing factors^8^, and studies such as eMERGE^9^ are developing a framework for integrating and returning polygenic risk score (PRS) results in healthcare settings. As PRSs approach clinical application, it is critical to establish a framework that promotes improved health for all individuals.

The clinical use of PRSs for risk prediction faces significant challenges, particularly in diverse populations. Differences in linkage disequilibrium patterns and allele frequencies arising from demographic history can lead to substantial variation in PRS distributions across populations. These differences do not necessarily reflect variation in disease risk, and careful standardization is needed to compare PRSs across individuals of diverse ancestry^10,11^. Furthermore, the predictive accuracy of PRSs developed from primarily European ancestry (EA) populations is substantially lower in non-European groups—especially among individuals of African ancestry^12–14^—with performance declining as the genetic distance between the training and target populations increases^15,16^. While ongoing efforts aim to improve performance by increasing the ancestral diversity of GWASs^3,17,18^ and by developing methods to account for diverse populations^19–24^, disparities in training sample sizes continue to limit prediction accuracy in underrepresented groups.

PRS prediction accuracy for disease traits is commonly described using metrics such as relative risk per standard deviation or closely related measures of discriminatory performance, such as AUC and C-statistics, none of which are sensitive to individuals’ background risks arising from various contextual factors. Relative-risk-based evaluations have often led to the conclusion that the clinical utility of current PRSs is lower in non-European and especially African-ancestry populations^12,25^. The clinical utility of a PRS for an individual, however, depends on many factors, involving complex interactions between age, sex, lifestyle factors, social determinants of health, genetic ancestry, and self-identified race/ethnicity. Baseline risks of many common diseases are strongly influenced by age, sex, and modifiable risk factors, and it has also been shown that PRS accuracy can vary across these participant characteristics and health-related contexts^26–28^. Importantly, there are well-documented differences in disease incidence rates across race and ethnic groups that cannot be explained by genetic factors alone. Social determinants, such as education, income, and healthcare access, can substantially impact treatment, prognosis, and downstream economic consequences following disease onset^29–34^. Thus, fully interpreting the impact of polygenic risk across diverse populations requires moving to more context-aware, life course-informed frameworks.

Absolute risk models provide ^35,36^ a natural framework to assess the added value of PRSs in conjunction with other established risk factors and have been applied to assess the potential utility of PRSs for risk-stratified screening ^5,37^ and targeted lifestyle interventions ^37,38^. However, the application of absolute risk models to evaluate PRS utility across diverse ancestry populations has been limited by several challenges. First, unlike relative risk and AUC, which provide convenient summary measures of model discrimination performance, there are no suitable metrics to summarize the impact of individual risk factors within integrated absolute risk models. There is also a lack of frameworks for developing absolute risk models specifically designed for diverse populations, such as those in the US, that jointly account for self-reported race/ethnicity and genetic ancestry, which are correlated but carry independent information. Moreover, because social and structural factors may more strongly influence disease outcomes (e.g., survival) than disease onset, there is a need to extend the absolute risk framework to incorporate post-diagnostic outcomes. In this article, we propose an absolute risk-based framework, together with a measure of variable importance, to evaluate the contextual utility of PRSs for stratifying disease risk and disease-specific mortality, while accounting for race/ethnicity, genetic ancestry, and other relevant population characteristics. We demonstrate the utility of our framework using data from the All of Us research program, focusing on breast (BRCA) and prostate (PRCA) cancers. Our findings demonstrate the limitations of interpreting PRSs without contextualizing risk within an absolute risk framework. The proposed framework can be generalized to incorporate information on contexts beyond race, ethnicity, and genetic ancestry, such as social determinants of health and lifestyle factors.

## Results

### Overview of Methods

An overview of the analysis and underlying methods is provided in **Fig. 1** and Briefly, we develop methods to calculate individualized absolute risks of future disease incidence and disease-specific mortality using information on PRS, age, genetic ancestry, self-identified race and ethnicity (SIRE) groups, and potentially other contextual variables. In developing models for breast and prostate cancer, we utilize established PRSs and their known associations across SIRE groups and use All of Us “training” data to model disease-risk variation across genetic ancestries. We allow the absolute-risk calculations to be calibrated with age-specific population incidence and post-incidence survival rates, stratified by SIRE groups, using data from the NCI-SEER cancer registry. Models were validated using held-out All of Us validation data and then used to project 5-year absolute risk and 10-year mortality for cancer-free participants across SIRE groups. We further introduce the Conditional Average Derivative Estimator (CADE), which quantifies the impact of PRS on the absolute risk of disease incidence or mortality within specific contexts, such as age strata and SIRE groups. Typically, the effect of continuous risk factors like PRS on absolute risks is highly non-linear and varies strongly with other risk factors. We define CADE to capture the gradient of the absolute-risk function with respect to the PRS, averaged over its population distribution.

**Figure 1.**
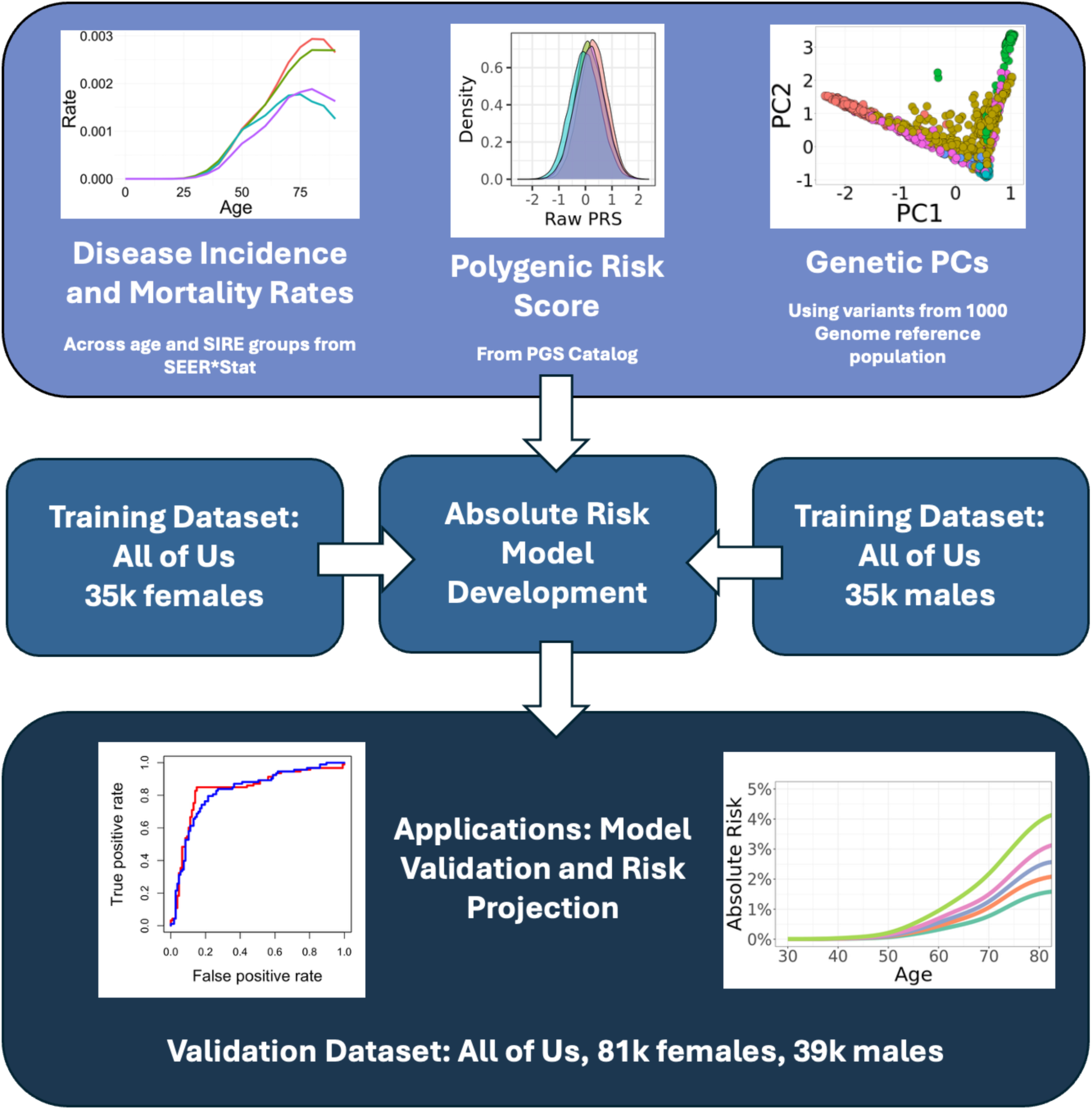
Absolute risk modeling framework for cancer incidence and cancer-specific mortality. The absolute risk model combines disease incidence rate information from SEER*Stat, polygenic risk scores from the PGS Catalog and reported associations across SIRE groups, and genetic PCs generated from a subset of variants from the 1000 Genomes reference population. The All of Us training subset is used to model cancer risk in relation to genetic PCs, after accounting for risk associated with PRS and SIRE groups. The All of Us validation subset is used to evaluate the discriminatory performance of alternative modes of risk evaluation using standardized PRS and absolute risk models. Finally, absolute risks for future cancer are projected for cancer-free individuals in the All of Us validation set. Cases in All of Us were defined by examining both electronic health records (EHR) and medical history survey responses.

### Development and Validation of Absolute-Risk Models for Breast and Prostate Cancers

We use All of Us training data (N=118,083 females, N=73,931 males) to model the risk of breast (**Supplementary Table 1**) and prostate cancers (**Supplementary Table 2**) in relation to established PRSs (PGS000004 and PGS003765), SIRE groups, and principal components of genetic ancestry. For both cancers, we observe a strong association (OR_BRCA_ = 1.44, 95% CI: 1.40-1.48; OR_PRCA_ = 2.02, 95% CI: 1.95-2.10) between the PRS and the risk of respective cancer among non-Hispanic White (NHW) individuals, and attenuated PRS effects for the other SIRE groups, with patterns generally consistent with those reported in prior studies^1,2,39–41^. We also observe significant variation in baseline cancer risk across the SIRE groups, with the baseline risk for other groups relative to NHW ranging from 0.10 to 0.49 for BRCA and from 0.45 to 0.77 for PRCA. The observed patterns are generally consistent with known variations in SEER incidence rates of these two cancers across the same groups. Notably, statistically significant associations with cancer risk remain for several genetic PCs after adjusting for SIRE groups for both cancers. The magnitude of association for some PCs is substantial: PC1 (OR per s.d. unit = 1.29) and PC4 (OR = 1.27) for BRCA, and PC2 (OR = 0.81) and PC4 (OR = 1.43) for PRCA. The final absolute risk model for breast cancer incidence incorporates the PRS [PGS000004], externally reported effect sizes across SIRE groups^1,39–41^ (**Supplementary Table 3**), and the top three most significant genetic principal components (PCs) selected based on All of Us training data analysis. The final model for prostate cancer similarly includes the PRS (PGS003765) and externally reported effect sizes across different SIRE groups (**Supplementary Table 3**), and the top three significant genetic PCs.

Validation analysis within the All of Us data suggests that the two models generally produce well-calibrated relative risks across risk deciles in populations where there were sufficient sample sizes for such evaluation (**Supplementary Fig. 1**). Further, evaluation of risk discrimination suggests that standardization of PRSs by genetic ancestry improves performance compared to standardization by SIRE categories (**Table 1**). For example, the number of breast cancer cases identified in the top 5% of PRS values increased by almost 25% (from 390 to 480) when the PRS was standardized by genetic ancestry rather than by SIRE groups. Further, the absolute risk model, which incorporates information on age-specific incidence rates by SIRE categories, improves discriminatory performance at both the high- and low-risk ends. The absolute risk model also shows a gain in discriminatory performance within age groups (**Supplementary Tables 4 and 5**), although the magnitude of improvement was smaller because variation in risk due to age does not contribute to the discriminatory performance of models in such analysis. While there are clear enrichments of cancer cases at various high-risk thresholds, a large proportion of the total number of cancer cases is missed by these thresholds. For example, only 28.7% and 41.8% of the total number of breast and prostate cancer cases, respectively, arise in the top 10% of the population identified by the respective absolute risk models. In contrast, low-risk thresholds provide strong assurance against the onset of these cancers.

**Table 1.** Number of cancer cases identified in the All of Us validation analysis at various risk thresholds defined by standardized PRS, individual-level standardized PRS, and the individual-level disease absolute risk model. Group-standardized PRS adjusts for SIRE-level mean and variance before ranking individuals across the population. Individual-level standardized PRS performs mean–variance adjustment based on principal components of genetic ancestry. The individual-level absolute risk model accounts for variation in PRS accuracy, distribution, and cancer incidence rates by age, genetic ancestry, and SIRE information. Models with higher discriminatory performance are expected to identify higher and lower numbers of cases at the top and bottom risk thresholds, respectively. PRS used for breast and prostate cancer are PGS000004 and PGS003765, respectively. SIRE: Self-identified race and ethnicity.

| Scoring Method | Breast Cancer (5,504 total cases) |  |  | Prostate Cancer (2,604 total cases) |  |  |
| --- | --- | --- | --- | --- | --- | --- |
|  | Top 5% | Top 10% | Top 20% | Top 5% | Top 10% | Top 20% |
| Group Standardized PRS | 390 | 743 | 1406 | 194 | 349 | 711 |
| Individual-level Standardized PRS | 482 | 893 | 1593 | 357 | 620 | 1030 |
| Individual-level Absolute Risk | 921 | 1577 | 2642 | 2642 | 1078 | 1656 |
|  | Bottom 5% | Bottom 10% | Bottom 20% | Bottom 5% | Bottom 10% | Bottom 20% |
| Group Standardized PRS | 167 | 356 | 770 | 49 | 127 | 283 |
| Individual-level Standardized PRS | 159 | 318 | 661 | 36 | 101 | 248 |
| Individual-level Absolute Risk | 5 | 14 | 68 | 1 | 2 | 4 |

### Projection of Absolute Risk of Cancer Onset and Mortality in Cancer-Free Individuals

Projections of the 5-year risk for the All of Us participants across age groups 45–50, 55–60, and 65–70 (**Fig. 2**) show that, for the same standardized PRS value, non-Hispanic White women tend to have higher projected breast cancer risk compared to other SIRE groups. In particular, we observe the strongest gradient in absolute risk due to variation in PRS values within the NHW population, driven by both a stronger relative risk associated with the PRS (see **Supplementary Table 3**) and a higher baseline risk in this group. The corresponding gradient in risk is similar for Asian women, but considerably lower for Black and Hispanic women. The CADE values (**Table 2**), which summarize these descriptive patterns, reveal that the utility of this PRS for absolute risk stratification of breast cancer in Black and Hispanic women is 50% or lower compared to NHW women.

**Figure 2.**
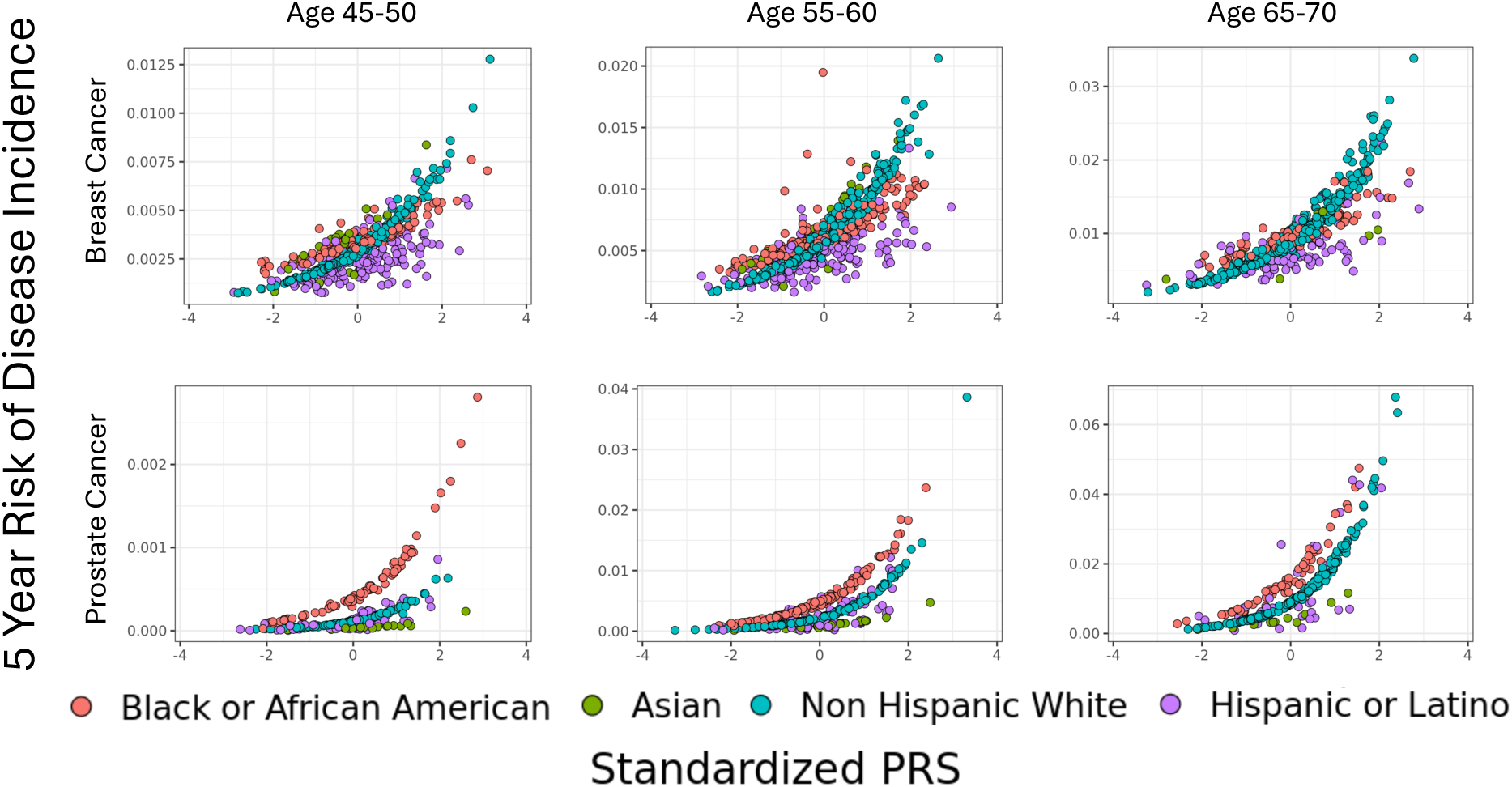
Projection of 5-year absolute risk of cancer incidence in relation to standardized PRS across age and SIRE groups for cancer-free individuals in the All of Us validation dataset. For each subplot, the X-axis shows PC-standardized PRS values for individuals in a specific category and the Y-axis shows the corresponding variation in 5-year risk. The absolute risk model accounts for variation in risk due to differences in PRS accuracy and age-specific baseline risk across SIRE categories and genetic ancestry.

**Table 2.**
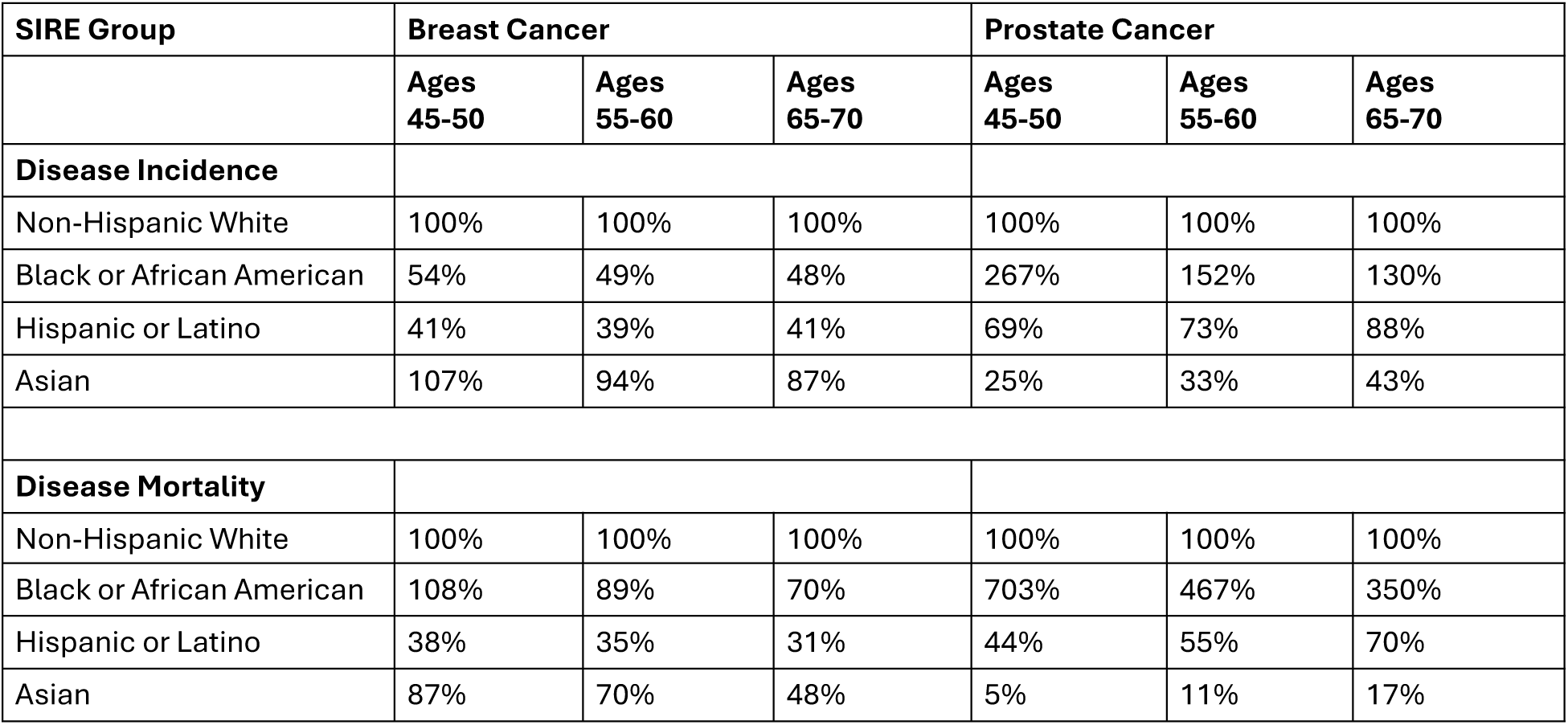
Relative CADE estimates for comparing PRS utility for absolute risk stratification in non-European population groups compared to European Americans. . The conditional average derivative estimate (CADE) summarizes the absolute-risk gradient associated with PRS averaged over the underlying population distribution of PRS. The CADE estimates are derived under an assumed log-linear model for risk, under which they can be approximated by the product of the log-relative risk associated with PRS in a specific context and the average population risk of the outcome (incidence or mortality) in the same context.

| SIRE Group | Breast Cancer |  |  | Prostate Cancer |  |  |
| --- | --- | --- | --- | --- | --- | --- |
|  | Ages<br>45-50 | Ages<br>55-60 | Ages<br>65-70 | Ages<br>45-50 | Ages<br>55-60 | Ages<br>65-70 |
| **Disease Incidence** |  |  |  |  |  |  |
| Non-Hispanic White | 100% | 100% | 100% | 100% | 100% | 100% |
| Black or African American | 54% | 49% | 48% | 267% | 152% | 130% |
| Hispanic or Latino | 41% | 39% | 41% | 69% | 73% | 88% |
| Asian | 107% | 94% | 87% | 25% | 33% | 43% |
| **Disease Mortality** |  |  |  |  |  |  |
| Non-Hispanic White | 100% | 100% | 100% | 100% | 100% | 100% |
| Black or African American | 108% | 89% | 70% | 703% | 467% | 350% |
| Hispanic or Latino | 38% | 35% | 31% | 44% | 55% | 70% |
| Asian | 87% | 70% | 48% | 5% | 11% | 17% |

For prostate cancer, the magnitude of absolute risks across PRS levels was highest for Black men, especially at younger ages (45-50 age group), reflecting the known high rate of prostate cancer in this group. The gradient in absolute risk associated with the PRS also appeared strongest for Black men, especially in the youngest age groups. The absolute risk levels and underlying gradients were lowest among Hispanic and Asian men compared with the other two groups. These patterns were reflected in the corresponding CADE values (**Table 2**). Compared to NHW men, the CADE values were considerably higher for black men (from 1.3 to 2.6-fold depending on age), somewhat lower for Hispanic men (from 0.9 to 0.7-fold), and substantially lower for Asian men (from 0.45 to 0.25-fold). The relative risks for the underlying PRS (**Supplementary Table 3**) were comparable across groups; thus, in this example, the pronounced differences in CADE values were mainly driven by differences in baseline rates.

We next projected the 10-year cause-specific mortality risk for breast cancer (women) and prostate cancer (men) in the All of Us validation sample (**Fig. 3**). In stark contrast to the patterns observed for breast cancer disease risk (**Fig. 2**), when mortality was considered, we observe that among women aged 45–50, Black women are projected to have consistently higher risk than women in the other groups, regardless of their standardized PRS values. This pattern reflects considerably worse survival rates for early-onset breast cancer in Black women observed in the SEER database, which are incorporated into our modeling framework, and are likely driven by a higher prevalence of hormone-negative breast cancer among young black women^42^. Within this age group, we observe that the CADE value of PRS utility for mortality risk (see **Table 2**) is fairly comparable for Black and White women, despite much lower relative risk for the PRS in the former group (OR per s.d. unit=1.26) compared to the latter (OR=1.6) (see **Supplementary Table 3**). With increasing age, the mortality gap between African American and non-Hispanic White women narrows, approaching overlap by ages 65–70, and correspondingly, the relative CADE value drops below 1.0. Finally, the CADE values reveal that, across all ages, the current PRS will have substantially lower utility for breast cancer mortality risk stratification for Hispanic women compared to all other groups.

**Figure 3.**
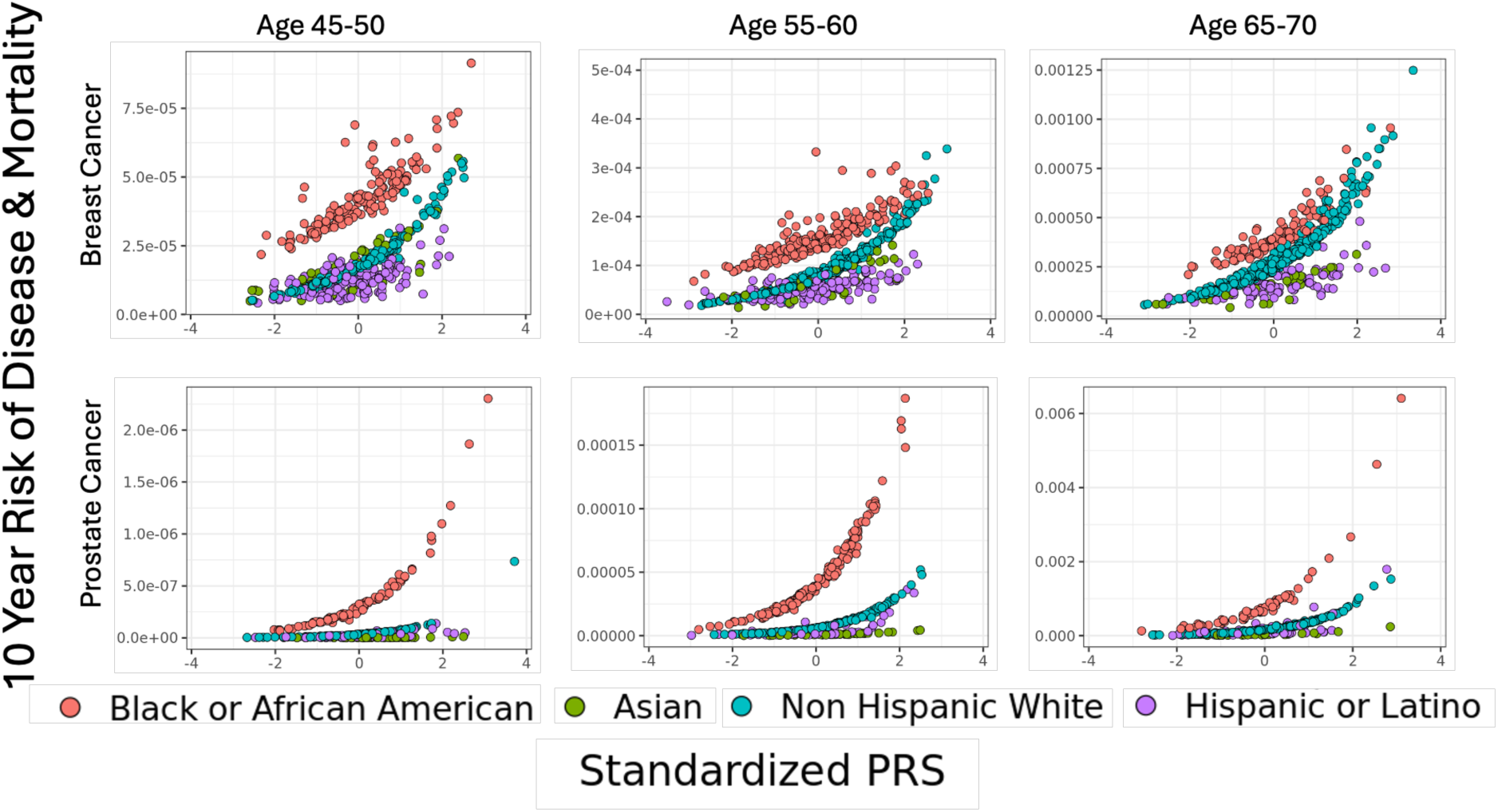
Projection of 10-year absolute risk of cancer incidence and cancer-specific mortality in relation to standardized PRS across age and SIRE groups for cancer-free individuals in the All of Us validation dataset. For each subplot, the X-axis shows PC-standardized PRS for individuals in a specific category and the Y-axis shows the corresponding variation in 10-year risk of cancer-specific mortality. The absolute risk model accounts for all factors influencing disease incidence as well as additional variability associated with mortality following cancer onset due to age at onset and SIRE groups.

For prostate cancer mortality, both the levels of absolute risk and the gradient in risk are consistently higher for Black men than for all other SIRE groups across PRS levels and age strata—a contrast to the incidence curves, where Black–White differences diminish at older ages (**Fig. 3**). This pattern reflects the poorer survival rates for Black men across age groups observed in the SEER database. CADE values for prostate cancer mortality risk for Black men were 3.5- to 7-fold higher compared to their NHW counterparts. The CADE values also reveal that the PRS will have minimal utility for risk stratification for prostate cancer mortality for Asian men compared to the other groups. This is again notable given that the relative risk for this prostate cancer PRS for Asian men (2.15) is quite comparable to other groups (2.04-2.32), and the pattern is driven by the low risk of incidence and relatively high survival rates in this group compared to the others.

Finally, we compared the SIRE group composition in the top 5% of individuals across the three scoring approaches (**Fig. 4**). When individuals are ranked by the standardized PRS, the composition of the highest-risk group reflects their composition in the underlying population—majority White, then Black and Hispanic, and then Asians representing the smallest group. When individuals are ranked based on absolute risk, the composition changes dramatically. For breast cancer, non-Hispanic White women formed the plurality or majority of individuals when the absolute risk of incidence was defined as high risk. In contrast, Black or African American women predominated in high-risk groups defined by mortality, particularly at younger ages. For prostate cancer, ranking by absolute risk led Black individuals to dominate the highest-risk group, nearing 100% when considering mortality at younger ages. On the other hand, the use of absolute risk of either incidence or mortality leads to the identification of very low numbers of Asian men in the highest-risk group.

**Figure 4.**
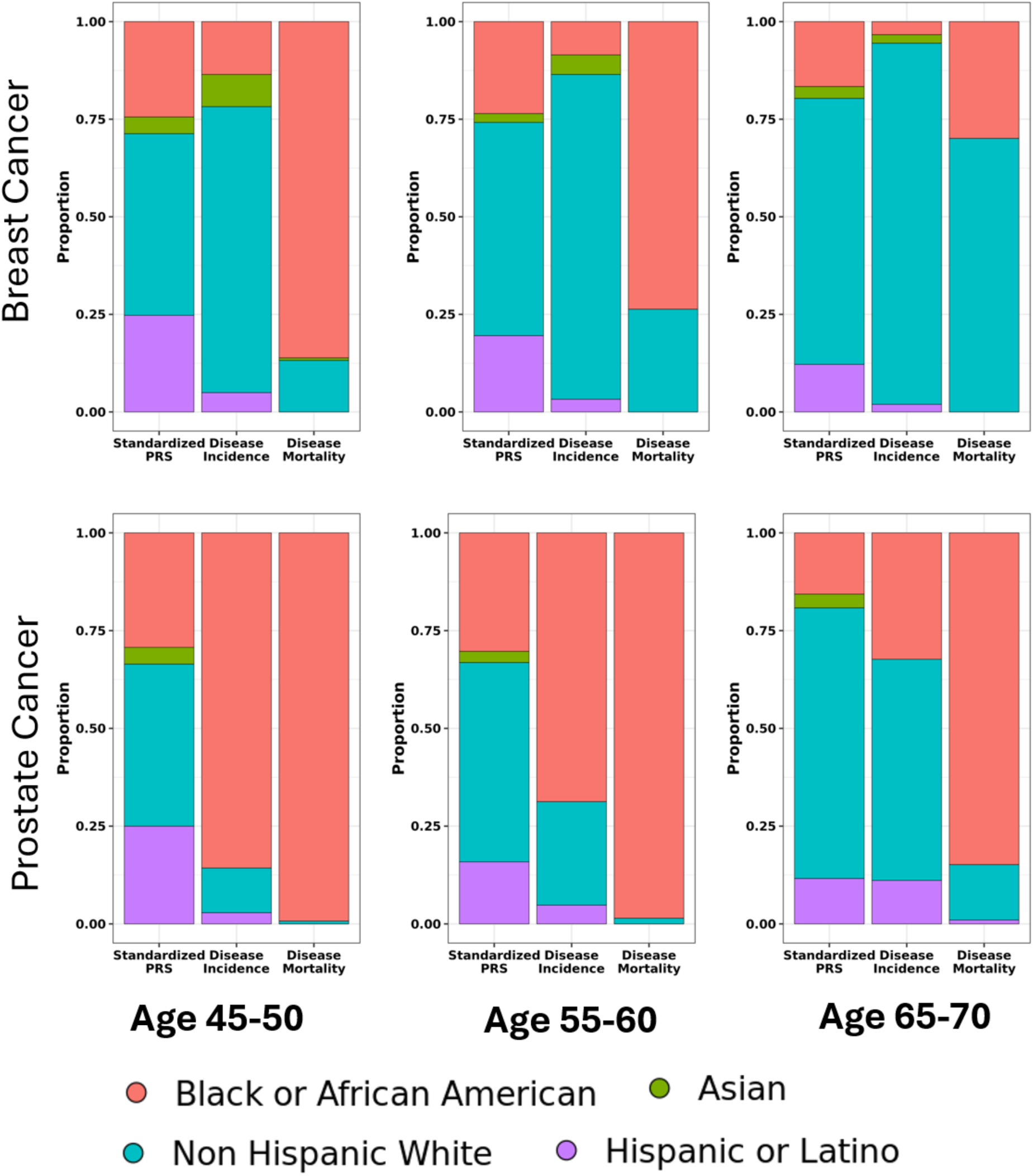
Distribution of SIRE groups among the top 5% highest-risk cancer-free individuals in the All of Us validation set. Individuals in different age categories were first ranked according to standardized PRS, 5-year absolute risk of cancer incidence, and 10-year risk of cancer-specific mortality. For each ranking, the proportions of individuals in the top 5% belonging to different SIRE categories were then evaluated.

## Discussion

In this article, we propose an absolute risk modeling framework and an associated measure of variable importance that can be used to compare PRS utility across individuals of diverse backgrounds, incorporating information on individualized genetic ancestry and known population-level variations in disease risks and mortality across self-identified groups. We used this framework to demonstrate that, while established PRSs for breast and prostate cancer—like many other complex traits—have the lowest relative risk of associations for African ancestry populations, these PRSs may have the greatest utility for absolute risk stratification in African American men for prostate cancer and African American young women for breast cancer. On the other hand, while the relative risks for the same PRSs are comparable between Asian and European Americans, their utility for absolute risk stratification in the former population may be fairly poor due to low baseline risks. Thus, while larger and more diverse GWASs are still needed to narrow the gap in PRS accuracy across diverse groups, assessment of the clinical utility of current PRSs in certain populations should be withheld on the basis of measures of relative risk (eg: OR) and AUC alone, without considering an absolute risk modeling framework.

Our findings also have implications for reporting genetically-informed risk assessments to individuals in diverse populations, such as those in the US, highlighting the need for careful consideration of the actual burden of disease, not only due to polygenic risk but also to societal, ancestral, and environmental contexts, as well as existing comorbidities. Studies have consistently shown that established PRSs achieve the largest magnitude of relative risk of association and discriminatory accuracy in European ancestry individuals^12–16,43^, with reductions of 50% or greater for African ancestry individuals. There are also distributional shifts in PRSs across diverse populations, which require careful standardization before applying a single threshold to define the high-risk group. In the eMERGE study^9^, which is developing standards for PRS reporting, individuals are classified as having high polygenic risk if they are in the top 5% of the PRS distribution after standardization using genetic ancestry principal components. While this standardization is important for removing differences in PRS distributions that reflect population structure, determining risk based on the PRS alone is inadequate, as it does not convey the differential burdens individuals face due to differences in risk factors and modifiers.

There are existing frameworks, such as iCARE^36^, and methodologies underlying popular tools, such as BCRAT^44^ and BOADICEA^45^, for developing individualized absolute risk models calibrated to population incidence rates. Development of population-calibrated absolute risk models for diverse populations, accounting for continuous genetic ancestry as well as commonly used population descriptors, which are correlated but carry independent information, requires additional considerations. We developed a modeling framework that allows genetic ancestry to influence PRS distribution, and both genetic ancestry and population descriptors to influence disease risk. Under this framework, we developed analytic formulae for model calibration using genomic reference datasets and population incidence rates (see **Methods**). We further extended the framework to estimate individualized risk of disease-specific mortality, accounting for factors associated with survival rates. In the future, the model could be extended to incorporate additional contextual factors, e.g., lifestyle factors and social determinants of health, which are known to influence disease incidence rates and/or prognoses.

In the past, PRS clinical utility across diverse ancestry populations has typically been reported using gradients of relative risk associated with the standardized PRS—commonly expressed as relative risk, hazard ratio, or odds ratio—which also have close one-to-one relationships with measures of discriminatory performance, such as the AUC statistic. Based on these measures, it is generally concluded that the clinical utility of existing PRSs is the poorest for African ancestry populations, resulting in concerns about exacerbating health inequities due to PRS implementation ^12^. However, application of the proposed framework for predicting breast and prostate cancer risk and mortality indicated that PRS utility in fact sometimes can be the highest for African American populations, due to elevated baseline risks associated with other factors. We note that accounting for differential survival rates following disease onset, even if they were not strongly related to the PRSs, can lead to very different perspectives on the life-course burden individuals face due to genetic susceptibility in the background of various contextual factors. In the future, evaluation of PRS utility should go beyond analysis of disease incidence and incorporate other endpoints, including mortality, disability-free life expectancy, and healthcare expenditures.

Our risk analysis of breast and prostate cancers has several limitations. Most importantly, in the absolute risk models for both breast and prostate cancers, we incorporate risk parameters associated with genetic principal components derived from analyses of the All of Us “training data.” In All of Us, disease diagnoses are reported using a combination of self-reported and longitudinal electronic health record (EHR) data. Because both self-reported diagnoses and EHR-based ascertainment may introduce measurement error in case–control determination, and because such error may itself be related to genetic ancestry, the association between certain genetic PCs and cancer risk observed in All of Us may be specific to this dataset. Although variations in disease risk and mortality rates by SIRE groups are well documented in population registries and cohort studies, future prospective studies of diverse individuals are needed to determine how genetic ancestry affects these rates, especially after accounting for SIRE group information. Another limitation is that we incorporate PRS association parameters only by SIRE group, even though PRS performance has been shown to degrade continuously as a function of the genetic distance between the training and test samples ^15^. However, our framework can be readily extended to calibrate the underlying relative risk parameters using continuous genetic ancestry information as it becomes more widely available. Finally, we develop the model for mortality for breast and prostate cancers, assuming the PRSs have negligible effects on survival following disease onset. While recent studies support this assumption for cancers and other common diseases^46–48^, it may not universally hold for other conditions, such as cardiovascular disease, where a PRS is associated with recurrent events following the first onset^49^. In the future, cohort studies of diverse ancestry individuals could be used to evaluate the impact of PRSs on cause-specific mortality directly.

In conclusion, we have developed a novel context- and life-course-aware framework to evaluate the clinical utility of PRSs for risk stratification across diverse populations. The application of this framework to breast and prostate cancer risk reveals clear limitations of commonly used approaches for PRS evaluation and PRS reporting across individuals of diverse ancestries. Adapting similar frameworks to other conditions can lead to more effective and equitable use of PRSs for risk-stratified prevention and improved health and mortality outcomes.

## Methods

### An Absolute Risk Model for Disease Incidence incorporating Age, PGS, Race/Ethnicity, and Genetic Ancestry Contexts

An overview of the modeling framework is provided in **Supplementary Figure 2**. We begin by establishing a model to calculate the absolute risk of a binary outcome *D*, such as disease incidence. For factors that can affect the absolute risk in this model, we consider the genetic variation associated with the disease, summarized by the *PRS*, the SIRE group *R*, and the genetic ancestry, *A*, as well as additional contexts, such as age, which we represent as *C*. Observe that *R* is a categorical variable. In contrast, *A*, which can be proxied by the genetic principal components, is continuous, while *C* can take many forms, such as age or social determinants of health. Furthermore, we assume that the *PRS* for individual *i* is given by *PRS_i_* = ***G***_***i***_*β*, where ***G***_***i***_ is the individual’s genotype and *β* are the weights of the genetic variants.

Now, we wish to model the individual-level absolute risk, or Pr(*D_i_* = 1| *R_i_*, *A_i_*, *C_i_*, *PRS_i_*). For simplicity, we assume a log-linear model, though this may be extended to other models of absolute risk, including logistic and time-to-event models. Assuming the log-linear model, for individual *i,* we have:

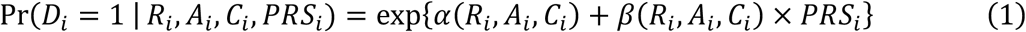

where *α*(*R_i_*, *A_i_*, *C_i_*) is the baseline risk, and *β*(*R_i_*, *A_i_*, *C_i_*) is the effect size of the *PRS* on the probability of disease.

It is important to consider potential data sources for estimating the different components of the models. First, the context-specific effect size of the PRS, denoted *β*(*R*, *A*, *C*), can be estimated by modeling interactions between the PRS and various contextual factors, such as self-identified race/ethnicity (R), genetic ancestry (A), and other covariates (C), using suitable study designs. Differential PRS effect sizes across SIRE groups have been reported for multiple traits and are increasingly cataloged in resources like the PGS Catalog^1,2,39–41^. Additionally, PRS performance has been shown to degrade with increasing genetic distance between the training and target populations^15^, highlighting the need for models that incorporate continuous ancestry measures. Ideally, *β*(*R*, *A*, *C*) it should be estimated using data from prospective cohort studies with incident cases, which provide unbiased effect size estimates. However, for rare diseases, the limited number of cases often precludes reliance on cohort data alone. In such settings, large-scale biobanks and linked EHR datasets have been used to calibrate PRS effects across contexts^26^. Still, accurately modeling context-dependent variation in PRS effects related to dynamic lifestyle and behavioral factors ultimately requires well-designed cohort studies with longitudinal follow-up^50^.

Estimation of the baseline risk, *α*(*R_i_*, *A_i_*, *C_i_*), is particularly sensitive to bias due to non-representative samples and incomplete follow-up. It generally cannot rely on case-control or convenience-based EHR cohorts. Later, we describe a strategy for using external population-based data to calibrate reliably *α*(*R_i_*, *A_i_*, *C_i_*). Thus, building absolute risk models across diverse populations may require estimating *β*(*R_i_*, *A_i_*, *C_i_*) and *α*(*R_i_*, *A_i_*, *C_i_*) using complementary data sources—balancing statistical precision against the risk of bias, depending on the parameter being estimated.

### A Model for the Distribution of Polygenic Risk Score

We must also establish a model for the distribution of polygenic risk scores in the underlying population across the various contexts under consideration. As PRSs are weighted averages of SNPs, we can model them as following a normal distribution, with models for the underlying mean and variance. In general, PRS distributions are strongly related to genetic ancestry^51^ and to certain environmental covariates due to population stratification^14^. In the following, for simplicity, we will assume the PRS follows a model in which its distribution depends only on genetic ancestry. Still, this model can be extended to incorporate additional contextual factors. We do assume that self-identified race and ethnicity are a poor proxy for genetic ancestry^43^ and that the PRS distribution is conditionally independent of SIRE information, given genetic ancestry.

Therefore, for an individual *i*, we can model their PRS conditional on their race *R*, genetic ancestry *A_i_*, and additional contexts *C_i_*, as a normal distribution with:

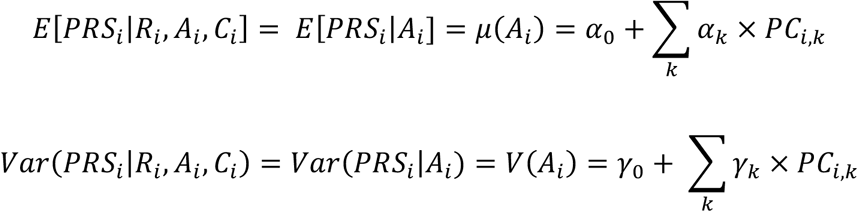

where the PCs are the genetic principal components for the individual, which have been used for individual-level mean and variance standardization in the same manner in current relative risk methodology ^9,10^.

### Calibration of Absolute Risk Model by Population Disease Incidence Rates

To calculate an individual’s absolute risk from their polygenic risk score (PRS) using Equation (1), we must also estimate the term *α*(*R_i_*, *A_i_*, *C_i_*), which reflects the baseline risk as a function of their self-identified race/ethnicity (R*i*), genetic ancestry (A*i*), and other contextual factors (C*i*). Ideally, *α*(*R_i_*, *A_i_*, *C_i_*) and the other model parameters would be estimated using data from prospective cohort studies that represent the target population. However, genetic studies are often based on convenient case-control samples, which have well-known limitations for estimating risk parameters associated with environmental and lifestyle factors. Cohorts like All of Us, which rely on electronic health records and self-reported outcomes, face similar challenges due to difficulties in identifying true incident cases and issues with incomplete follow-up.

In contrast, high-quality cohort studies like the UK Biobank, with linkage to national health registries and well-defined follow-up, are expected to provide more reliable estimates for associations with environmental and behavioral factors. However, even these population-based cohorts may not yield unbiased estimates of absolute disease incidence due to volunteer bias, limiting their use for generalizable population-level risk prediction. To overcome these limitations, it is often necessary to supplement model estimations with population-based disease incidence rates from national surveys and registries. In cancer epidemiology, this approach has a long history—calibration techniques using data from registries such as SEER have been widely used to construct absolute risk models ^36,52^. In the context of multi-ancestry modeling, we now propose extending these calibration techniques where the risk model may be based on genetic ancestry. Still, registry data are only available at the level of self-identified race/ethnicity groups.

In our modelling, we consider a specific decomposition of the baseline risk in the form

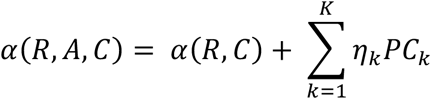

where we will estimate *α*(*R*, *C*) through calibration with population incidence rates of the disease, Pr(*D* = 1 | *R*, *C*) = *p_R,C_*, from external sources like cancer registries, and assume additional data sources are available to estimate the disease-association parameters for the genetic PCs, *η*_1_,….. *η_K_*, after adjusting for (*R*, *C*). In our application, we used All of Us data for the estimation of such association parameters.

For calibration, we solve the equations

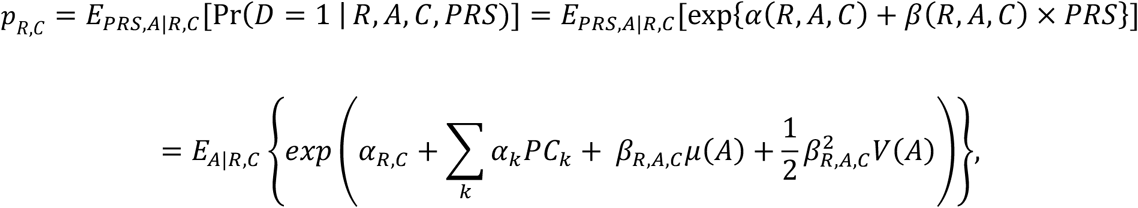

where the last expression is derived under the assumption for normal distribution of PRS and the mean-variance specification of it by genetic ancestry as described above. Further, the outside expectation involves computing the expectation with respect to the ancestry variable, ie. genetic PCs, conditional on race and context. In our example, where *C* = *age*, we assume the distribution of the PCs only depends on SIRE groups, and we use and we use half of the All of Us training data of the opposite sex, males for breast cancer and females for prostate cancer, to calculate respective averages for the estimation of the underlying expectations. Using this, and the estimates for disease incidence by SIRE group and contexts, we can solve for *α_R,C_* and then fit the final absolute risk model as follows:

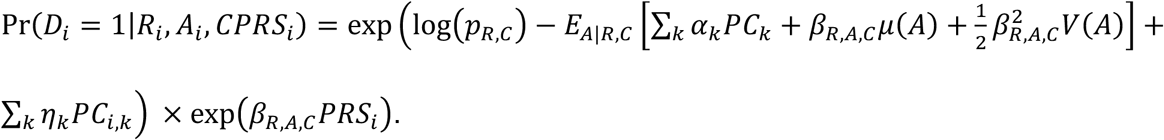

For the absolute risk model applied to the All of Us cohort, we assume *β_R,A,C_* = *β_R_* and use their estimates from publicly available sources (**Supplementary Table 3**). Further, we obtained population incidence rates for cancer by SIRE and age groups from SEER*Stat.

### Model for Absolute Risk of Disease-specific Mortality

We can further extend the model to consider the impact of polygenic burden of a disease on disease-specific mortality. We specify the relationship of the risk of disease-specific mortality to the PRS of the underlying disease and the contextual factors as:

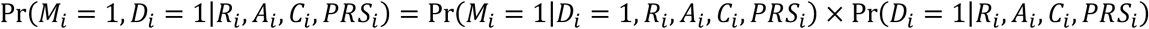

where *M_i_* is the indicator of mortality following disease incidence.

As previous research has shown little association between PRS and mortality after accounting for disease incidence^53,54^, and current data sources do not have estimates for the effect of genetic ancestry on mortality, for our application, we will simplify the above model to:

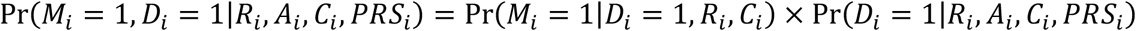

Observe that we have previously described how to build the second half of this model after calibrating with the population incidence rate. Additionally, the model for mortality after disease incidence can be built based on prospective data of patient populations and baseline context variables, such as SIRE, social determinants of health, or lifestyle factors. For our applications, we utilize estimates of mortality rates after cancer incidence stratified by age and SIRE groups using data from SEER.

### Conditional Average Derivative Estimator

PRS utility for disease risk prediction, within and across populations, are often reported using standardized relative risk, area under the curve, or c-statistics, none of which take into account the absolute risk of diseases. For a binary predictor with two levels, *Z* = 0 or 1, a useful measure for risk-stratification and the effect of intervention is the risk-difference parameter, which is the difference in the absolute risk of disease between the two categories. For continuous biomarkers like PRSs, such risk-difference parameters could be defined based on certain categorizations, but this requires subjective decision-making regarding the number and placement of the categories. Motivated by average derivative estimator for summarizing association under smoothed regression model^55^, below we define a novel context-dependent summary measure for risk-stratification for a continuous predictor (*X*) like the PRS.

Let *A_c_*(*x*) be the absolute risk of the disease, as estimated from an underlying “model”, at the level of the predictor *X* = *x* and given context *C* = *c*. Let 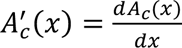 be the gradient of the risk function at *X* = *x*, and define a measure of conditional average derivative estimator (CADE), in the context, *C* = *c*, as

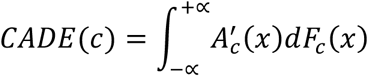

where *F_c_*(*x*) is the distribution function for *X* conditional on *C* = *c*. Intuitively, the CADE can be viewed as risk gradient parameters over small changes in *X* integrated over its underlying distribution in the population. In the special case where the risk function has a log-linear form, such as *A_c_*(*x*) = exp(*α_c_* + *β_c_x*), where *α_c_* and *β_c_* are context specific baseline risk and risk-parameters, then one can derive

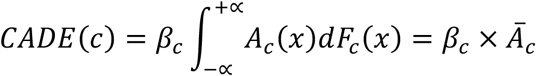

where *A̅*_*c*_ is the average population risk for the risk in the context *C* = *c*. This special case immediately shows that the CADE is a useful measure for predictive utility that combines information on log-relative-risk parameters and the average absolute risk for the population. One can also define tail-based measures of CADE that focus on the upper and lower tails of risk distribution at specific thresholds, say *x_L_* and *x_U_*, as

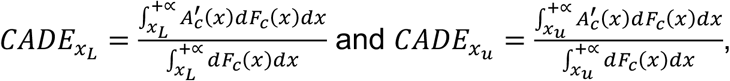

which, under log-linear model, can again be derived as the product of log-relative-risk parameters and the average risks over the restricted range of the distribution of *X*. We present relative CADE values associated with the PRSs for both risk and mortality of the two cancers (breast for women and prostate for men) to compare the risk-stratification utility of these PRSs across population groups defined by race and ethnicity. The relative CADE value for context *C* = *c* compared to a baseline context *C* = *c*_0_ can be calculated as

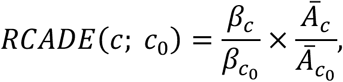

which involves the product of the ratio of log-RR of PRS across the two populations and the ratio of the population average-risk across the two populations, with the later quantity corresponding to relative-risk (RR) of the outcome associated with one context compared to another.

### Data Sources

#### All of Us

We illustrate the application of our framework using data available in the All of Us research program. All of Us is a growing research program overseen by the National Institutes of Health, consisting of over half a million diverse American participants, with detailed survey, biomedical, genetic, and medical record data for a subset of its participants^56^. Our study population consists of those with whole genome sequencing and electronic health records (EHR) data, and define disease cases as anyone who included a record of the ICD-9 or ICD-10 codes for the cancer in the condition or observation domain within the EHR, or anyone who reported having a personal medical history of the condition in the medical history survey (**Supplementary Fig. 3**). We further filter the data to include only individuals whose self-identified race and ethnicity included Non-Hispanic White, Non-Hispanic Black, Non-Hispanic Asian, or Hispanic. In the application, we use about half of the individuals from All of Us to examine the risk of breast and prostate cancers associated with the PRS, self-identified race/ethnicity information, and genetic principal components. We projected the absolute risk of the cancers based on the proposed framework on the remaining half of the All of Us sample and carried out validation analysis based on the observed case-control status of these participants (**Supplementary Fig. 3**).

Specifically, after restricting the population to individuals who have both genetic and EHR data, and filtering for only individuals who self-identify as Non-Hispanic White, Non-Hispanic black, Non-Hispanic Asian, or Hispanic, our final data includes 118,083 females (8,119 breast cancer cases) and 73,931 males (4,982 prostate cancer cases). Approximately 35,000 individuals were sub-sampled for model training, to determine the effect sizes of the genetic ancestry PCs on the disease status. The remaining individuals were used for risk projection and validation (discrimination) analysis. The validation datasets consisted of 81,083 female participants (5,504 cases) for breast cancer, and 38,931 male participants (2,640 cases) for prostate cancer.

#### SEER*Stat

To obtain the population level disease incidence and mortality rates, we use the software provided by the National Cancer Institute’s Surveillance, Epidemiology, and End Results (SEER) program, which summarizes the data from SEER and other cancer registries to provide statistics for one year disease incidence and mortality^57^. For our analysis, we use the cancer incidence data available from 12 registries from 1992 to 2020, which we match by the site and morphology site, and then stratify by 5-year age groups and race and origin recode^58^. Similarly, for the mortality data, we use the incidence-based mortality data available from 12 registries from 1992 to 2020, which we match by site and morphology site, and again stratify by 5-year age at death groups and race and origin recode^59^. As SEER*Stat’s rates are reported for Non-Hispanic White, Non-Hispanic Black, Non-Hispanic Asian, and Hispanic, these are the SIRE groups we use in our analyses.

### Data Analysis

#### Multi-ancestry Principal Components for All of Us and 1000G

To model the relationship between the PRS and the principal component in our validation dataset, we first calculate the principal component (PC) loadings for the individuals from the 1000 Genomes project^60^, which we then use to calculate the PCs for both the 1000 Genomes and the All of Us validation set individuals. Specifically, we use PLINK2 using the –pca flag and use the HapMap3 SNPs to construct the PCs on the full 1000 Genomes population^61^. We then use PLINK2 to project both the 1000 Genomes and All of Us individuals onto the loadings to calculate the specific PCs.

#### Polygenic Risk Scores

We selected the 313 variant breast cancer PRS by Mavaddat et al.^1^, the PRS currently used in the BOADICEA model for breast cancer^45^, and the 451 variant prostate cancer PRS by Wang et al, a recent score developed in a multi-ancestry population^2^. The weights used for the analysis are taken from the PGS Catalog, and the odds ratios for association of the PRS with their respective cancers, stratified by racial and ethnic groups, were obtained from the PGS Catalog and the Wang et al. publication respectively. All variant effects were harmonized to the All of Us whole genome sequencing data using the function harmonise_data from the TwoSampleMR package.

#### Model Fitting and Validation

First, we divide our dataset into males and females and further divide each dataset into training and testing. We use a little under half of the individuals from the population of interest (female for breast cancer and male for prostate cancer) to explore association between breast cancer risk and genetics principal components, sampling the individuals from each SIRE group separately to ensure similar population distributions. We further use the individuals of the opposite sex as the “reference sample” to develop the mean and variance regression models for the PRS in relation to the principal components, and these models for PRS distributions were subsequently used for absolute risk calibrations within SIRE groups based on the methods described above. We also used these models to generate “standardized” PRS so that they are comparable across individuals of different ancestral backgrounds.

We used data from the training sample to fit logistic regression models for disease status as a function of the PRS, SIRE group, and genetic principal components simultaneously, with further adjustment for income and highest level of education as potential confounders. We incorporated interaction terms between PRS and SIRE groups, as our final models for absolute risk estimation incorporated SIRE group specific effect sizes for the PRS for both cancers. Estimates of the effect size for the genetic principal components from this model were then incorporated into the absolute risk model development for the population underlying the All of Us study. The associations of cancer risks with the PRS, SIRE group, and PRS by SIRE group interactions were used to explore the consistency of the data from what is known about these effects in published literature. Finally, we used individuals from the “testing sample” for evaluating the distribution of absolute risk of the cancers within and between different SIRE groups and evaluating the discriminatory ability of the model in identifying observed cancer cases compared to using the standardized PRS without accounting for additional contexts.

For the All of Us women and men in the validation samples, we also projected 10-year risks of cause-specific mortality for breast and prostate cancer, respectively, in relation to the PRS, SIRE groups and genetic ancestry. To calculate such absolute risk for an individual, we first found the probability of that individual developing the disease in the first *x* years, as taken from the absolute risk of disease incidence model, and then multiplied those values by the probability of dying of the disease in the next 10 − *x* years, as obtained from SEER*Stat, taking into account the shift in the age group over time. We compared the distribution of individuals of different SIRE groups among people who were considered at the highest 5% risk according to three criteria: (1) the standardized polygenic risk score, (2) the absolute risk of disease incidence, and (3) the absolute risk of disease-specific mortality.

## Acknowledgement

Research for this study was supported by NIH grant U01HG011719 (M.F., P.N, N.C), U01CA261339 (L.K, J.W) R01CA241410 (J.W.), R01HG013137 (N.C.) and R01CA228198 (D.L, N.C), R01CA228918 (D.H.). We gratefully acknowledge *All of Us* participants for their contributions, without whom this research would not have been possible. We also thank the National Institutes of Health’s *All of Us* Research Program for making available the participant data examined in this study

## Author’s Contribution

M.F led the development of the methods, conducted the data analysis, and drafted the manuscript. N.C conceptualized and supervised the study, and jointly drafted the manuscript with M.F., L.K., D.H., J.W. and P.N. All reviewed drafts of the manuscript and contributed to revisions.

## Data Availability

This study used data from the All of Us Research Program’s Controlled Tier Dataset 7, available to authorized users on the Researcher Workbench. Access can be obtained from https://researchallofus.org/. Cancer incidence and mortality rates are publicly available through the National Cancer Institute’s Surveillance, Epidemiology, and End Results (SEER) Program (https://seer.cancer.gov/data-software/documentation/seerstat/). Data was accessed using SEER*Stat software November 2023 Submission

## Code Availability

The code is available on Github at https://github.com/Fumm95/absolute-risk-and-cade.

