## Supplemental Tables and Figures for "Contextualizing the Utility of Polygenic Risk Scores using Absolute Risk Models in Diverse Ancestry Populations"

**Supplementary Figure 1. Calibration of the predicted and observed relative risks for breast and prostate cancer in the held-out validation dataset of All of Us.**

**Supplementary Figure 2. Absolute risk models for disease incidence and mortality and sources of information used for model building in our analysis.**

**Supplementary Figure 3. The number of female (Panel A) and male participants (Panel B) with EHR and Genetic Data in All of Us, and corresponding breast and prostate cancer cases identified from different sources.**

**Supplementary Table 1. Risk of breast cancer in relationship to PRS, SIRE groups and genetic ancestry in the All of Us training data.**

**Supplementary Table 2. Risk of prostate cancer in relation to PRS, SIRE groups, and genetic ancestry in the All of Us training data.**

**Supplementary Table 3.** **Comparison of performance of breast and prostate cancer PRSs in All of Us compared to those reported in external studies.**

**Supplementary Table 4.** **Number of breast cancer cases identified in the All of Us validation analysis within different age groups at various risk thresholds defined by the standardized PRS, individual-level standardized PRS, and individual-level disease absolute risk model.**

**Supplementary Table 5.** **Number of prostate cancer cases identified in the All of Us validation analysis within different age groups at various risk thresholds defined by the standardized PRS, individual-level standardized PRS, and individual-level disease absolute risk model.**

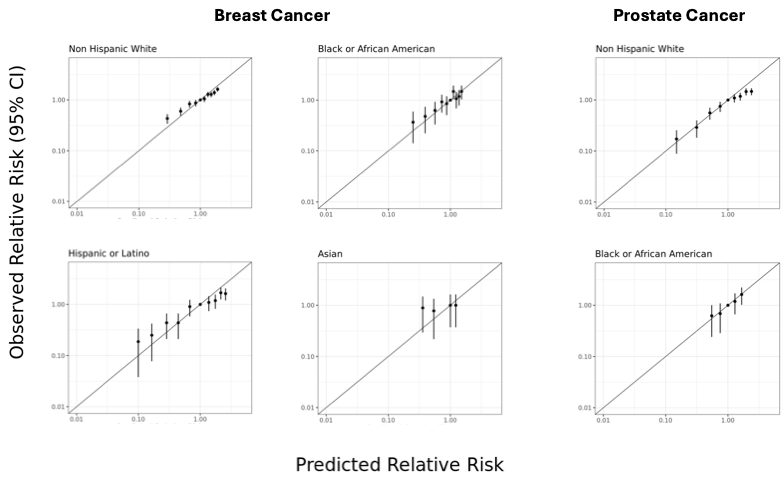

**Supplementary Figure 1. Calibration of the predicted and observed relative risks for breast and prostate cancer in the held-out validation dataset of All of Us.** Calibrations were attempted to be done with respect to risk deciles, but results are not shown for deciles that had too few observed cases (N ≤ 5). For prostate cancer, results are only shown for Non-Hispanic White and Black populations due to the small number of total cases (N ≤ 26) in the other two groups.

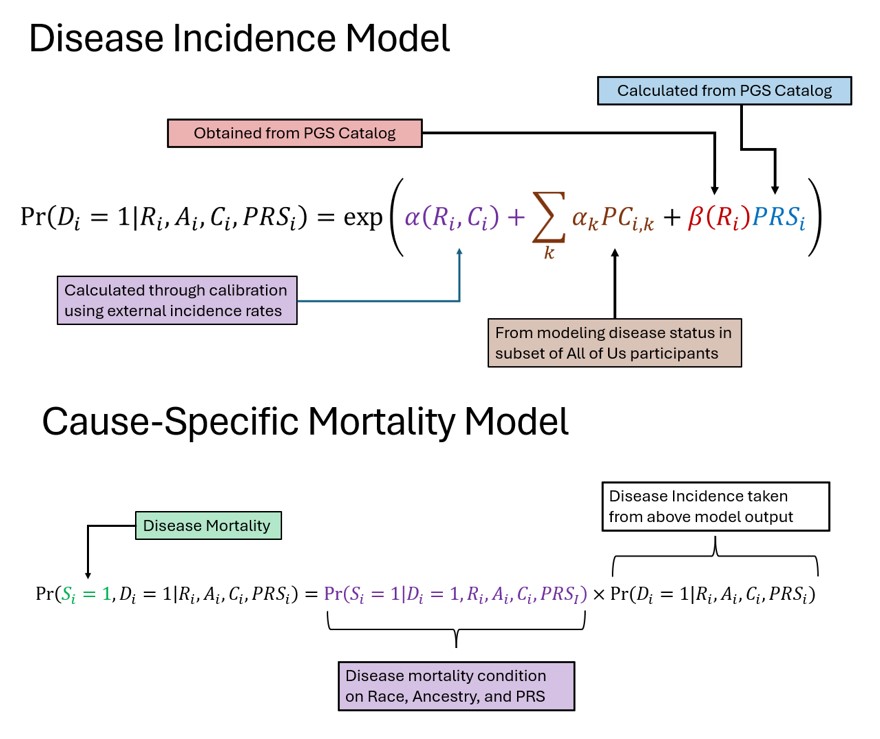

**Supplementary Figure 2. Absolute risk models for disease incidence and mortality and sources of information used for model building in our analysis.** To build the absolute risk model for disease incidence, we use established polygenic risk scores from the PGS Catalog alongside with their published effect sizes in different SIRE groups and use All of Us training data to model the association of cancer risk with genetic PCs after accounting for PGS and SIRE groups. The model for absolute risk of cancer onset is further calibrate using information on population incidence rates available by SIRE groups from the SEER database. Finally, the model for cancer survival following onset included only effect of age-at-onset and SIRE groups, information on which were also obtained from the SEER.

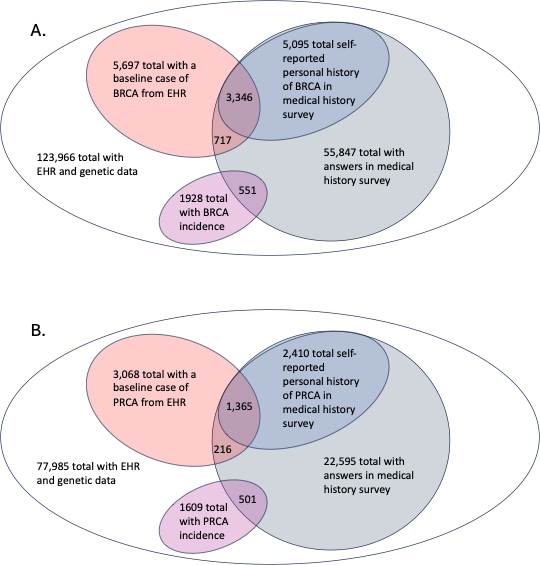

**Supplementary Figure 3. The number of female (Panel A) and male participants (Panel B) with EHR and Genetic Data in All of Us, and corresponding breast and prostate cancer cases identified from different sources.**

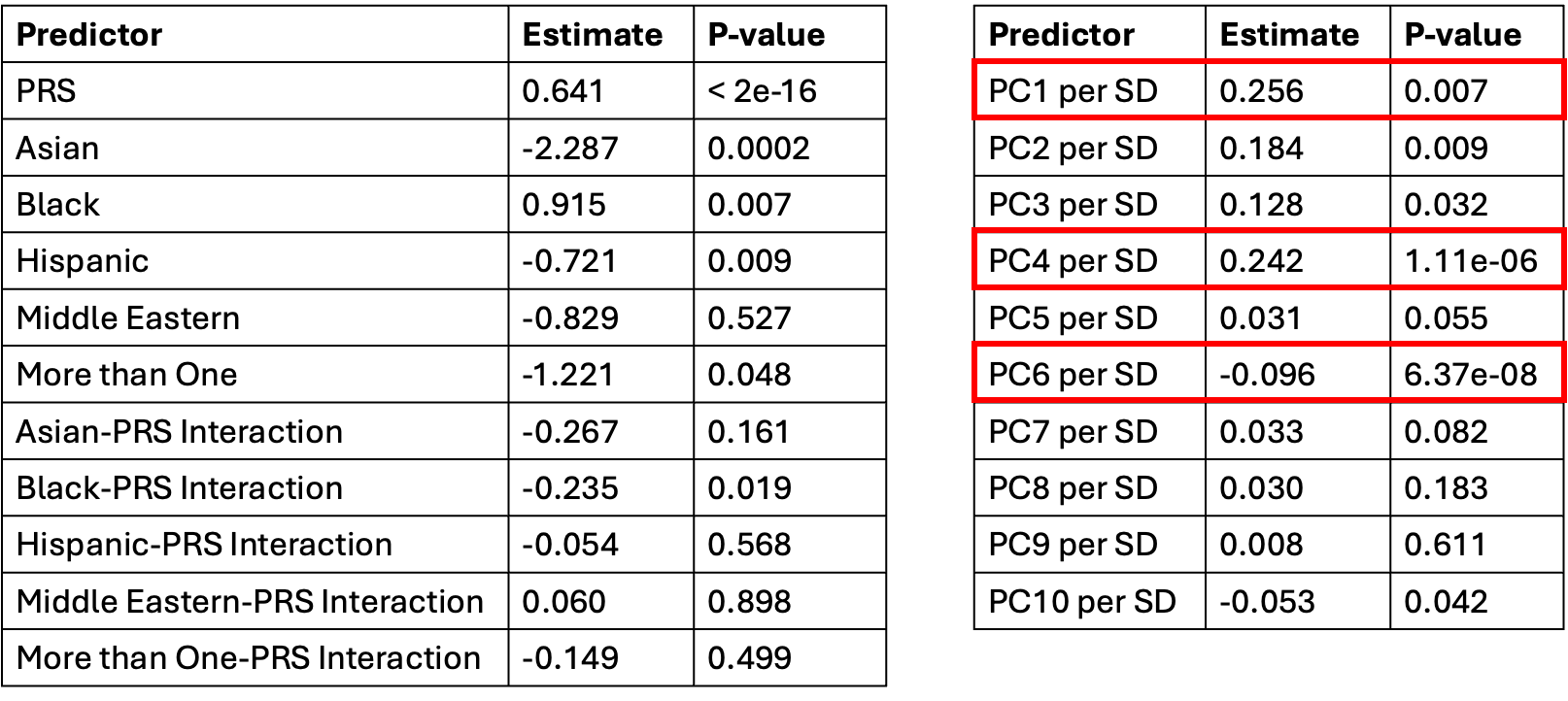

**Supplementary Table 1. Risk of breast cancer in relation to PRS, SIRE groups, and genetic ancestry in the All of Us training data.** The original PRS for breast cancer included 313 SNPs [PGSID: PGS000004], among which 298 were matched with the variants in All of Us, and was mean-variance standardized using genetic PCs. The genetic PCs for the All of Us participants were obtained based on SNP weights derived from 1000 Genome individuals.

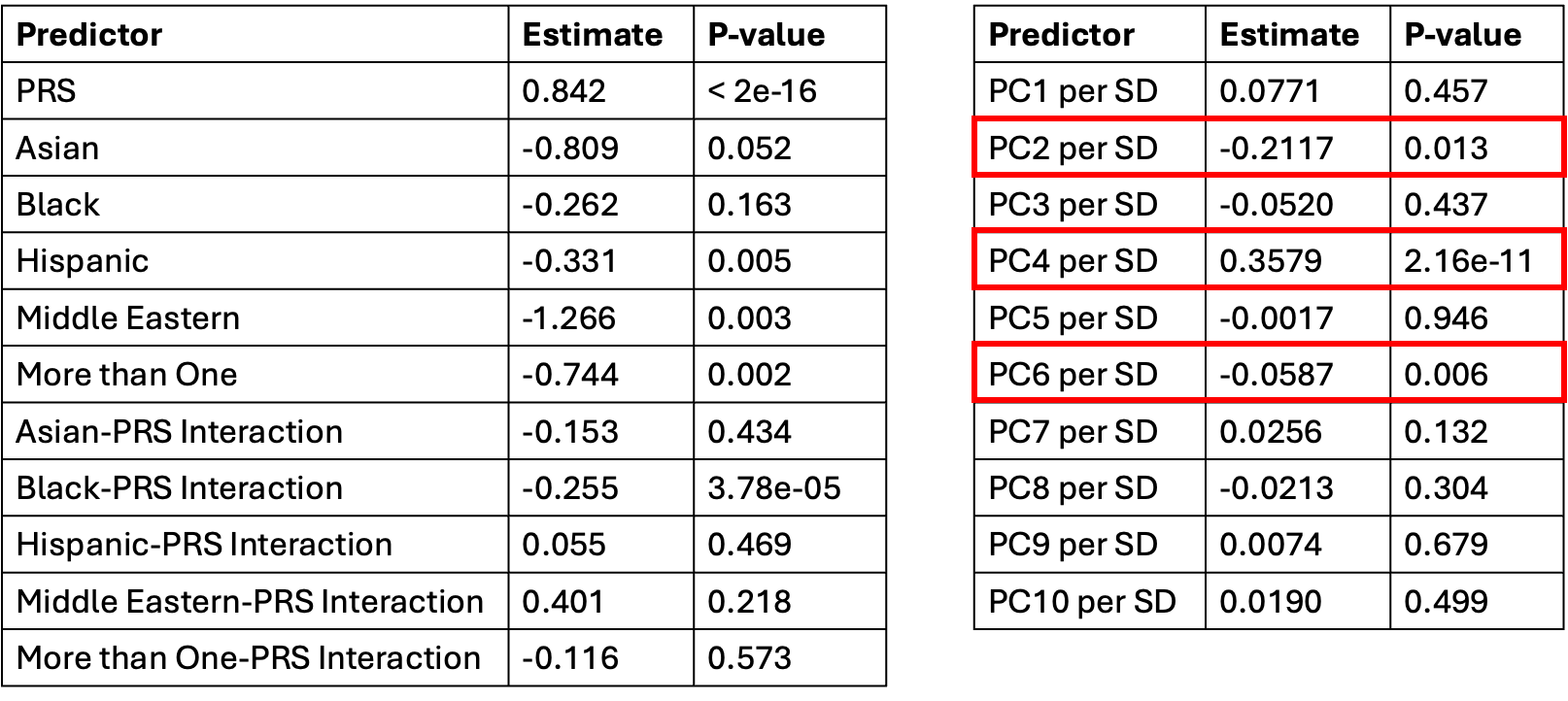

**Supplementary Table 2. Risk of prostate cancer in relation to PRS, SIRE groups, and genetic ancestry in the All of Us training data.** The original PRS for prostate cancer included 451 SNPs [PGSID: PGS003765], among which 386 were matched with the variants in All of Us, and was mean-variance standardized using genetic PCs. The genetic PCs for the All of Us participants were obtained based on SNP weights derived from 1000 Genome individuals.

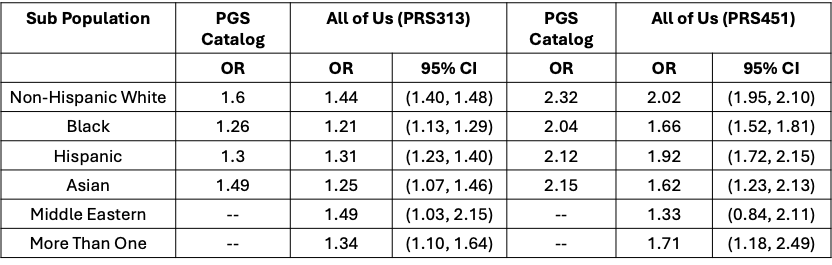

**Supplementary Table 3.** **Comparison of associations of breast and prostate cancer PRSs in All of Us compared to those reported in external studies.** For fair comparison with available external results, in this analysis, the PRS was not adjusted a priori for genetic PCs. The odds-ratios are presented per s.d. unit of PRS within each population group. For the final model building, we incorporated the odds-ratios reported in the PGS catalog.

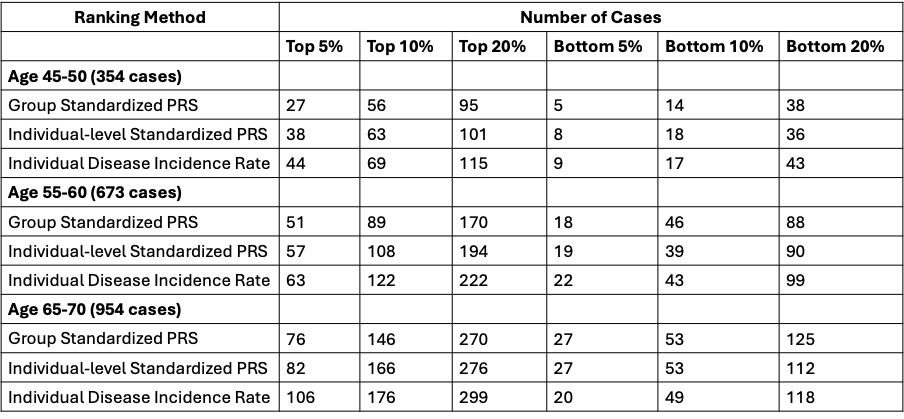

**Supplementary Table 4.** **Number of breast cancer cases identified in the All of Us validation analysis within different age groups at various risk thresholds defined by the standardized PRS, individual-level standardized PRS, and individual-level disease absolute risk model.** Group-standardized PRS adjusts for SIRE-level mean and variance before ranking individuals across the population. Individual-level standardized PRS performs mean–variance adjustment based on principal components of the genetic ancestry. The individual-level absolute risk model accounts for variations in PRS accuracy, distribution, and cancer incidence rates by age, genetic ancestry, and SIRE information. Models with higher discriminatory performance are expected to identify higher and lower numbers of cases at the top and bottom risk thresholds, respectively. PRS used for breast cancer is PGS000004. SIRE: Self-identified race and ethnicity.

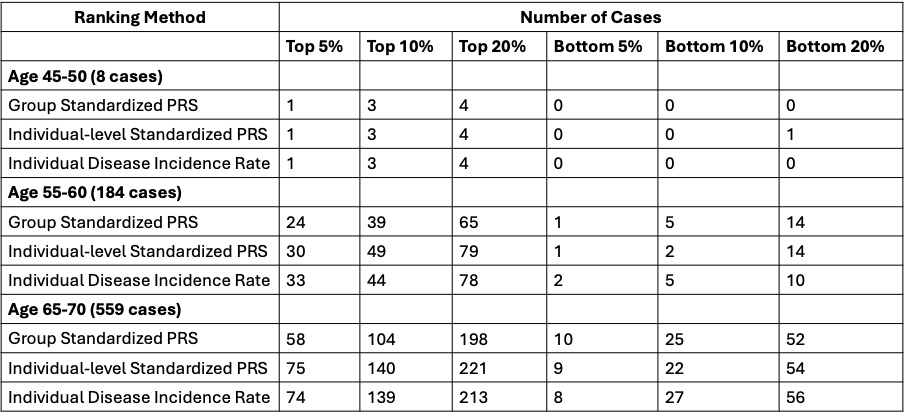

**Supplementary Table 5.** **Number of prostate cancer cases identified in the All of Us validation analysis within different age groups at various risk thresholds defined by the standardized PRS, individual-level standardized PRS, and individual-level disease absolute risk model.** Group-standardized PRS adjusts for SIRE-level mean and variance before ranking individuals across the population. Individual-level standardized PRS performs mean–variance adjustment based on principal components of the genetic ancestry. The individual-level absolute risk model accounts for variations in PRS accuracy, distribution, and cancer incidence rates by age, genetic ancestry, and SIRE information. Models with higher discriminatory performance are expected to identify higher and lower numbers of cases at the top and bottom risk thresholds, respectively. PRS used for prostate cancer is PGS003765. SIRE: Self-identified race and ethnicity.
